# Stacking Ensemble Learning-based Models Enabling Accurate Diagnosis of Cardiac Amyloidosis using SPECT/CT:an International and Multicentre Study

**DOI:** 10.1101/2025.06.22.25330039

**Authors:** Qu Mo, Jing Cui, Shengguo Jia, Ying Zhang, Yi Xiao, Caiguang Liu, Chunqing Zhou, Clemens P Spielvogel, Raffaella Calabretta, Weihua Zhou, Ke Cao, Marcus Hacker, Xiang Li, Min Zhao

**Affiliations:** Department of Nuclear Medicine, Xiangya Hospital, Central South University, Changsha 410008, China; Department of Nuclear Medicine, Third Xiangya Hospital, Central South University, Changsha 410013, China; Department of Nuclear Medicine, Fuwai Central China Cardiovascular Hospital, Henan Provincial People’s Hospital Heart Center, Zhengzhou 451464,China; Siemens Healthineers Ltd., Shanghai, China; Department of Biomedical Imaging and Image-Guided Therapy, Division of Nuclear Medicine, Medical University of Vienna, Vienna, Austria; Department of Applied Computing, Michigan Technological University, Houghton, MI, USA; Department of Oncology, Third Xiangya Hospital, Central South University, Changsha 410013, China

**Keywords:** Cardiac amyloidosis, Technetium pyrophosphate, Diagnosis, Radiomics, Machine learning

## Abstract

**PURPOSE:** Cardiac amyloidosis (CA), a life-threatening infiltrative cardiomyopathy, can be non-invasively diagnosed using [^99m^Tc]Tc-bisphosphonate SPECT/CT. However, subjective visual interpretation risks diagnostic inaccuracies. We developed and validated a machine learning (ML) framework leveraging SPECT/CT radiomics to automate CA detection.

**METHODS:** This retrospective multicenter study analyzed 290 patients of suspected CA who underwent [^99m^Tc]Tc-PYP or [^99m^Tc]Tc-DPD SPECT/CT. Radiomic features were extracted from co-registered SPECT and CT images, harmonized via intra-class correlation and Pearson correlation filtering, and optimized through LASSO regression. A stacking ensemble model incorporating support vector machine (SVM), random forest (RF), gradient boosting decision tree (GBDT), and adaptive boosting (AdaBoost) classifiers was constructed. The model was validated using an internal validation set (n = 54) and two external test set (n = 54 and n = 58).Model performance was evaluated using the area under the receiver operating characteristic curve (AUC), calibration, and decision curve analysis (DCA). Feature importance was interpreted using SHapley Additive exPlanations (SHAP) values.

**RESULTS:** Of 290 patients, 117 (40.3%) had CA. The stacking radiomics model attained AUCs of 0.871, 0.824, and 0.839 in the validation, test 1, and test 2 cohorts, respectively, significantly outperforming the clinical model (AUC 0.546 in validation set, *P*<0.05). DCA demonstrated superior net benefit over the clinical model across relevant thresholds, and SHAP analysis highlighted wavelet-transformed first-order and texture features as key predictors.

**CONCLUSION:** A stacking ML model with SPECT/CT radiomics improves CA diagnosis, showing strong generalizability across varied imaging protocols and populations and highlighting its potential as a decision-support tool.

## INTRODUCTION

Cardiac amyloidosis (CA) is a progressive disorder characterized by extracellular deposition of misfolded protein fibrils in myocardial tissue, leading to potentially fatal complications including heart failure and conduction abnormalities[1]. Despite advancements in diagnostic techniques, CA remains challenging to diagnose due to its heterogeneous and nonspecific clinical presentation, often resulting in diagnostic delays or misclassification[2].

While endomyocardial biopsy (EMB) remains the diagnostic gold standard, its invasive nature and associated risks have prompted the development of non-invasive alternatives [2]. [^99m^Tc]Tc-bisphosphonate scintigraphy has emerged as a reliable non-invasive diagnostic modality for transthyretin cardiac amyloidosis (ATTR-CA), particularly when combined with immunoglobulin light chain screening to exclude amyloid light chain cardiac amyloidosis (AL-CA) [3]. However, despite the advancements, the visual assessment method is inherently subjective, with accuracy heavily reliant on the physician’s experience[4]. Moreover, visual scoring has limitations in capturing the heterogeneity of diseases and its ability to reflect subtle disease information is limited, while being less effective in identifying AL-CA [5, 6].

Artificial intelligence(AI) has shown increasing promise for the diagnosis and prognostic assessment of CA[7]. In bone scintigraphy.Delbarre et al. trained a convolutional neural network on 3,048 images, achieving 99% sensitivity and 96% specificity versus expert visual reads [8]. Speilvogel et al. then implemented an automated image-standardization pipeline followed by deep learning(DL) to identify Perugini score ≥ 2 uptake, achieving an internal cross-validation AUC of 1.000 and external AUCs of 0.925-1.000 [9].

Hybrid SPECT/CT offers the distinct advantage of precise anatomical localization via co-registered images, which is especially valuable in early-stage disease when regional myocardial uptake may be mistaken for blood pool activity. Recently, a DL method was developed for fully automated volumetric quantification of [^99m^Tc]Tc-PYP using segmentation of co-registered anatomical structures from CT attenuation maps[4]. Despite these advances, AI -driven SPECT/CT applications for comprehensive CA diagnosis remain unexplored. This study aims to develop and validate an ensemble machine learning (ML) model based on SPECT/CT images for automated detection of CA, providing clinicians with a more reliable diagnostic tool.

## MATERIALS AND METHODS

### Study population

Data were retrospectively collected from 290 patients with suspected CA who underwent [^99m^Tc]Tc-bisphosphonate SPECT/CT imaging at three centers between February 2021 and February 2024. The training set and internal validation set from the center 1 (Xiangya Hospital of Central South University) consisted of 124 and 54 patients, respectively. The external test sets from the second center (Fuwai Central China Cardiovascular Hospital) and the third center (Vienna General Hospital, Medical University of Vienna) comprised 54 and 58 patients, respectively. Patients from Centers 1 and 2 received [^99m^Tc]Tc-PYP, while patients from Center 3 were administered [^99m^Tc]Tc-DPD as the imaging agent. A flowchart of patient enrollment is shown in ***Figure 1***. This study was granted ethical approval by the institutional review boards of Xiangya Hospital, Central South University (No. 202310202), Fuwai Central China Cardiovascular Hospital (Ethics Review No. 202303), and the General Hospital of Vienna (EK1557/2020). It was carried out in accordance with the principles of the 1964 Declaration of Helsinki. Owing to the retrospective study design, the Institutional Review Boards of all participating institutions formally waived the requirement for informed consent.

**Figure 1.**
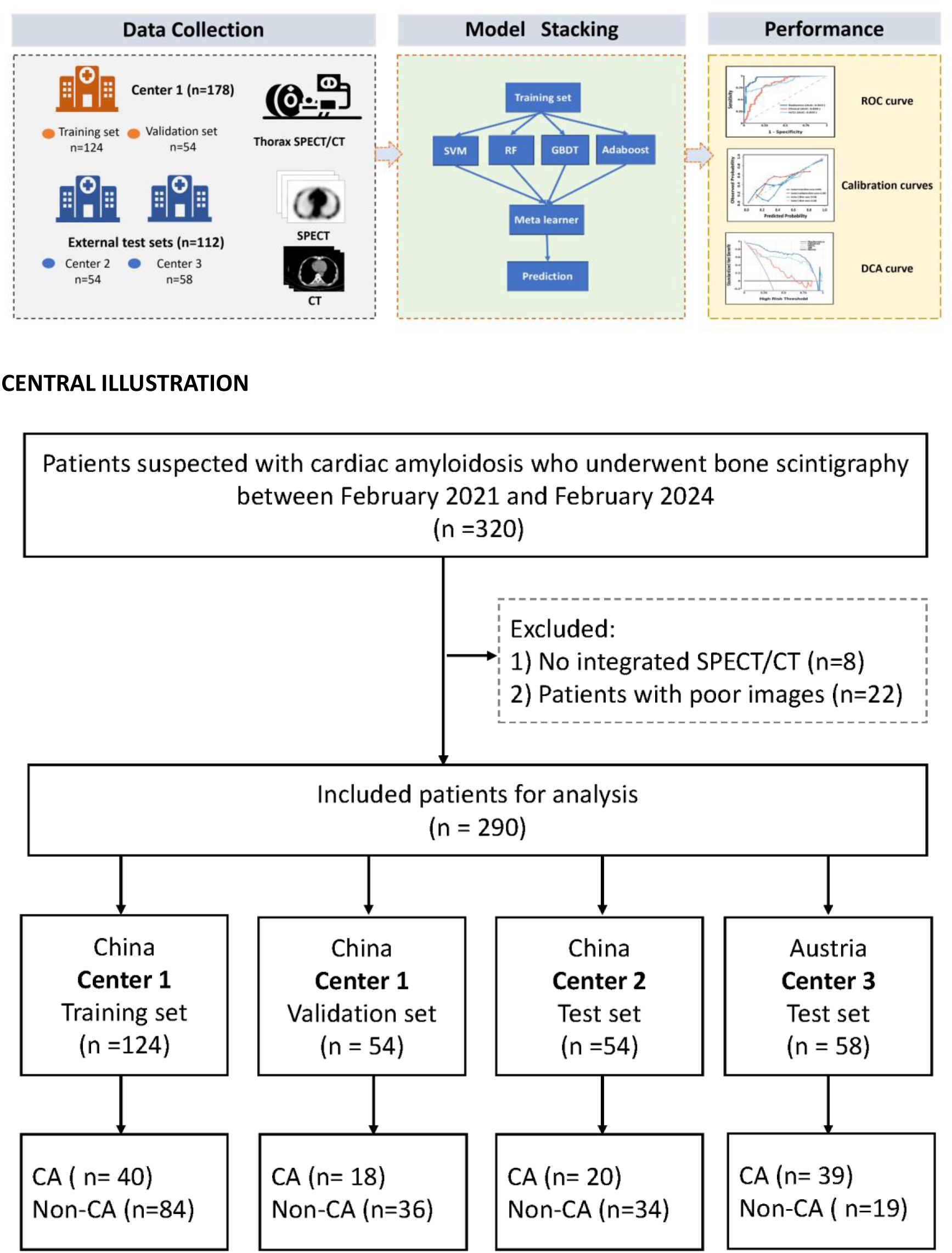
Flowchart of study cohort enrollment.

### Diagnosis of CA

Patients underwent a comprehensive diagnostic workup, which included cardiac imaging tests such as electrocardiogram (ECG), echocardiography, cardiac magnetic resonance imaging (CMR), and [^99m^Tc]Tc -biphosphonate scintigraphy, laboratory tests such as serum N-terminal prohormone of brain natriuretic peptide (NT-proBNP) and estimated glomerular filtration rate (eGFR), as well as myocardial or non-myocardial biopsy. The diagnosis of CA was made according to current guidelines[3, 10, 11]: CA was defined to be present if (1) [^99m^Tc]Tc-bisphosphonate imaging strongly suggested cardiac uptake in the absence of monoclonal gammopathy, the diagnosis was ATTR-CA. or (2) histological evidence of amyloid infiltration was found in endocardial or extracardiac biopsy tissues and cardiac imaging strongly suggested CA, with the presence of monoclonal immunoglobulin or free light chains in blood or urine, or monoclonal plasma cells/B cells in bone marrow, AL-CA was diagnosed or (3) other types of CA were confirmed by immunohistochemistry or mass spectrometry.

### SPECT/CT Imaging protocols

At Center 1&2, patients received an intravenous injection of 370-740 MBq of [^99m^Tc]Tc-PYP, followed by chest planar imaging 1 hour later and SPECT/CT of the thorax 3 hours post-injection. At Center3, patients were administered an intravenous injection of 700 MBq of [^99m^Tc]Tc-DPD, followed by whole-body planar imaging and chest SPECT/CT scan 3 hours later. CT scans were performed before SPECT acquisition to correct for radiation scatter and attenuation, and the SPECT data were reconstructed using the respective algorithms provided by each manufacturer. Detailed information on imaging vectors and acquisition protocols is provided in ***Supplementary Table 1***.

### Image analysis

Cardiac tracer uptake was assessed according to the Perugini grading system, where Grade 0 indicates no cardiac uptake, Grade 1 indicates mild cardiac uptake (lower than bone), and Grades 2 or 3 indicate intense cardiac uptake (equal to or greater than bone). Grades 0-1 were considered negative, while Grades 2-3 were considered positive[3, 9]. In each center, visual scoring was evaluated by two experienced (≥5 years) nuclear medicine physicians who were blinded to patient information. In cases where the visual scoring is ambiguous, to prevent disputes, consultation with a more senior physician is conducted to reach a consensus. A circular region of interest (ROI) was placed over the heart and mirrored to the contralateral chest (avoiding areas such as the sternum and stomach). The average counts within the ROI were calculated to obtain the heart-to-contralateral lung (H/CL) ratio. A 1-hour H/CL ratio ≥ 1.5 was considered positive, and a 3-hour H/CL ratio ≥ 1.3 was considered positive [3].

### Segmentation, radiomics feature extraction and selection

The volume of interest (VOI) for the heart contour was automatically segmented by the Siemens software MM RT Image Suite[12] (***Supplementary*** Figure 1), and subsequently reviewed and manually adjusted as needed by two experienced nuclear medicine physicians (Q.M., M.Z.),Radiomic features were extracted from both SPECT and CT images using PyRadiomics package (https://github.com/Radiomics/pyradiomics)[13], which is compliant with the Imaging Biomarker Standardization Initiative(IBSI). The extracted features can be divided into four sets: (1) first-order statistics; (2) shape-based features extracted from 2-D regions of interest or 3-D VOIs; (3) texture features including gray-level co-occurrence matrix (GLCM), gray-level dependence matrix (GLDM), gray-level run length matrix (GLRLM), gray-level run length matrix(GLRLM), gray-level sizezone matrix (GLSZM) and neighboring gray-level dependence matrix (NGTDM); (4) higher-order features using Laplacian of Gaussian (LoG) filter and wavelet transform filter with all possible combinations of high (H) or low (L) pass filter in each of the three dimensions (HHH, HHL, HLH, LHH, LLL, LLH, LHL, HLL).

All data from Center 1 were randomly split into a training set (70%) and an internal validation set (30%).Data from Centers 2 and 3 served as external testing sets. Optimal feature selection involved four sequential steps: (1) Z-score normalization of all features; (2) feature reproducibility was assessed via intra-class correlation coefficient (ICC), retaining only those with ICC > 0.75[16]; (3) Pearson correlation coefficient (PCC) analysis was performed, and when the PCC between any two features in the training set exceeded 0.90, one feature was removed to reduce redundancy[17]; and (4) LASSO regression with 1-standard error and 10-fold cross-validation was applied to avoid overfitting[18]. The radiomics workflow is shown in the ***Central Illustration***.

### Development and validation of the models

Independent predictive factors for CA in the training set were selected using univariate and multivariate logistic regression analysis. All significant clinical factors identified in the univariate analysis were included in the subsequent multivariate analysis. A clinical model was established based on the independent predictive factors. Based on the selected radiomic features, radiomics models were constructed using various ML algorithms, including support vector machine (SVM), random forest (RF), gradient boosting decision Tree (GBDT), adaptive boosting (Adaboost), and the stacking ensemble method. The stacking model utilized SVM, RF, GBDT, and Adaboost as primary learners (first level), with RF serving as the meta-learner (second level). ***Supplementary*** Figure 2 illustrates the development process of the stacking radiomics model.

By employing metrics such as area under the curve (AUC), accuracy(ACC), sensitivity(SEN), specificity(SPE), positive predictive value (PPV), and negative predictive value (NPV), we compared the performance among different models. while the clinical utility of the models in both training and validation cohorts was evaluated through decision curve analysis (DCA). Shapley Additive Explanations (SHAP) were employed to interpret the ML model by quantifying the contribution of each feature to the model’s predictions[19].

### Statistical analysis

All statistical analyses were performed using SPSS (version 27.0), R (version 4.4.1, (www.Rproject.org), and Python (version 3.10). Continuous data were represented using mean ± standard deviation (SD) or median (with interquartile range), depending on their distribution. When comparing continuous variables, either the t-test or the Mann-Whitney U test was used. Categorical data were analyzed by the chi-square test or Fisher’s exact test, and were presented as numbers (percentages). NT-proBNP levels were logarithmically transformed prior to inclusion in the logistic regression model. Univariate and multivariate regression analyses were conducted to identify risk factors. Multivariable logistic regression analyses were performed on variables with a P value < 0.1 from the univariate analysis. Model performance was assessed by plotting ROC curves, differences in AUCs between models were statistically compared using the DeLong test. A P value of less than 0.05 was considered statistically significant.

## RESULTS

### Patient characteristics

At the end of the diagnostic work-up, CA was diagnosed in 58 cases (32.6% of the whole population; 38 cases of ATTR-CA, 15 cases of AL-CA, and 5 cases of other types of CA) at Center 1, 20 cases (37.0% of the whole population; 3 cases of ATTR-CA, 13 cases of AL-CA, and 4 cases of other types of CA) at Center 2, and 39 cases (67.2% of the whole population; 35 cases of ATTR-CA, 4 cases of AL-CA) at Center 3. The clinical characteristics of the patients are summarized in ***Table 1***. Among these three centers, CA patients had significantly higher levels of NT-proBNP compared to non-CA patients. Furthermore, the H/CL ratio and visual score of CA patients were significantly higher than those of non-CA patients (both P<0.05).

**Table 1.**
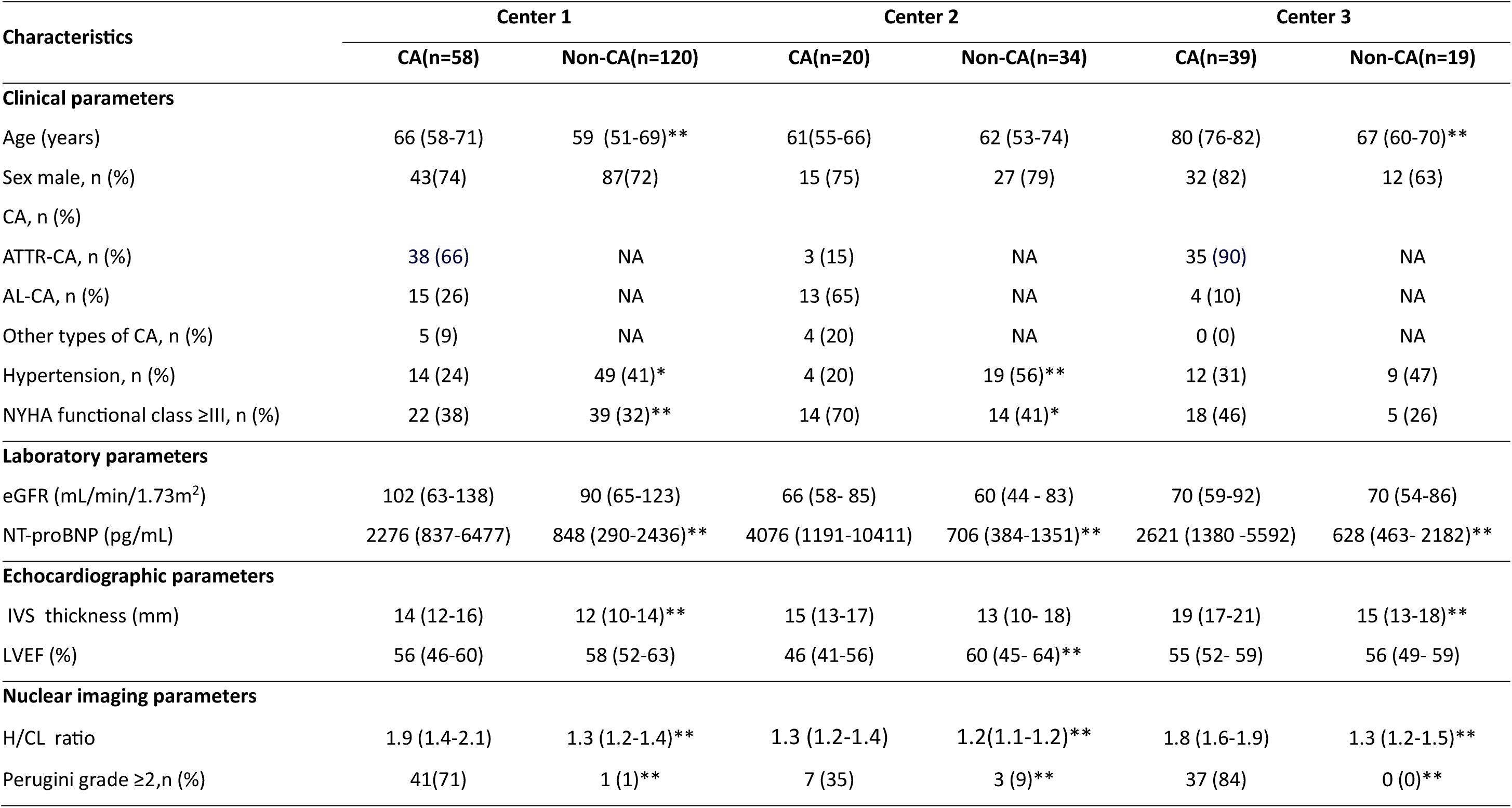
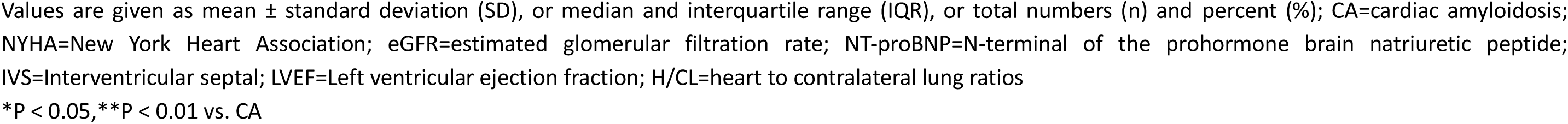
Characteristics of the patients in different cohorts.

| Characteristics | Center 1 |  | Center 2 |  | Center 3 |  |
| --- | --- | --- | --- | --- | --- | --- |
|  | CA(n=58) | Non-CA(n=120) | CA(n=20) | Non-CA(n=34) | CA(n=39) | Non-CA(n=19) |
| **Clinical parameters** |  |  |  |  |  |  |
| Age (years) | 66 (58-71) | 59 (51-69)** | 61(55-66) | 62 (53-74) | 80 (76-82) | 67 (60-70)** |
| Sex male, n (%) | 43(74) | 87(72) | 15 (75) | 27 (79) | 32 (82) | 12 (63) |
| CA, n (%) |  |  |  |  |  |  |
| ATTR-CA, n (%) | 38 (66) | NA | 3 (15) | NA | 35 (90) | NA |
| AL-CA, n (%) | 15 (26) | NA | 13 (65) | NA | 4 (10) | NA |
| Other types of CA, n (%) | 5 (9) | NA | 4 (20) | NA | 0 (0) | NA |
| Hypertension, n (%) | 14 (24) | 49 (41)* | 4 (20) | 19 (56)** | 12 (31) | 9 (47) |
| NYHA functional class ≥III, n (%) | 22 (38) | 39 (32)** | 14 (70) | 14 (41)* | 18 (46) | 5 (26) |
| **Laboratory parameters** |  |  |  |  |  |  |
| eGFR (mL/min/1.73m<sup>2</sup>) | 102 (63-138) | 90 (65-123) | 66 (58- 85) | 60 (44 - 83) | 70 (59-92) | 70 (54-86) |
| NT-proBNP (pg/mL) | 2276 (837-6477) | 848 (290-2436)** | 4076 (1191-10411) | 706 (384-1351)** | 2621 (1380 -5592) | 628 (463- 2182)** |
| **Echocardiographic parameters** |  |  |  |  |  |  |
| IVS thickness (mm) | 14 (12-16) | 12 (10-14)** | 15 (13-17) | 13 (10- 18) | 19 (17-21) | 15 (13-18)** |
| LVEF (%) | 56 (46-60) | 58 (52-63) | 46 (41-56) | 60 (45- 64)** | 55 (52- 59) | 56 (49- 59) |
| **Nuclear imaging parameters** |  |  |  |  |  |  |
| H/CL ratio | 1.9 (1.4-2.1) | 1.3 (1.2-1.4)** | 1.3 (1.2-1.4) | 1.2(1.1-1.2)** | 1.8 (1.6-1.9) | 1.3 (1.2-1.5)** |
| Perugini grade ≥2,n (%) | 41(71) | 1 (1)** | 7 (35) | 3 (9)** | 37 (84) | 0 (0)** |

Values are given as mean ± standard deviation (SD), or median and interquartile range (IQR), or total numbers (n) and percent (%); CA=cardiac amyloidosis; NYHA=New York Heart Association; eGFR=estimated glomerular filtration rate; NT-proBNP=N-terminal of the prohormone brain natriuretic peptide; IVS=Interventricular septal; LVEF=Left ventricular ejection fraction; H/CL=heart to contralateral lung ratios \*P < 0.05, \*\*P < 0.01 vs. CA

### Feature Selection

A total of 1888 features were extracted,from the CT and SPECT images. 376 features with high consistency were selected after-dimensionality reduction by ICC and PCC analysis, 9 key features were ultimately selected for the radiomics model. Among them, 4 features were extracted from SPECT images: Glszm_GrayLevel_NonUniformity, Log-sigma-3-0-mm-3D_ngtdm_Contrast, Wavelet-LLH_firstorder_Median, and Wavelet-LHH_glszm_GrayLevel_NonUniformity_Normalized. The remaining 5 features were derived from CT images: Firstorder_Entropy, Wavelet-LLH_firstorder_10Percentile, Wavelet-LLH_glszm_SizeZone_NonUniformity_Normalized, Wavelet-LLL_firstorder_90Percentile, and Wavelet-LLL_gldm_Dependence_Entropy.

### Univariate and multivariate analysis

The univariate analysis revealed that age, lgNT-proBNP, and Interventricular septal (IVS) thickness derived from echocardiography were associated with CA. Subsequently, the significant variables from the univariate analysis (age, lgNT-proBNP, IVS thickness) were included in the multivariate logistic regression analysis. In multivariable analysis, lgNT-proBNP (OR: 4.63, 95% CI: 2.09 - 10.29, p < 0.001) and IVS thickness (OR: 1.23, 95% CI: 1.07 - 1.40, p = 0.003) were independent predictors of CA (***Supplementary Table 2***). A predictive model constructed based on clinical parameters was then used to predict CA.

### Model Performance for Differentiating CA and Non-CA

We evaluated the diagnostic performance of radiomics models based on five ML algorithms. As shown in ***Figure 2***, the stacking ensemble algorithm model achieved the highest AUCs on the training set and on the Center 2 and Center 3 test sets, with slightly lower performance than SVM on the Center 1-validation set. Despite the lack of statistical significance, the stacking ensemble algorithm model exhibited smaller AUC fluctuations across datasets, suggesting relative robustness. The calibration curves for stacking ensemble model indicated strong agreement between predicted probabilities and observed outcomes across datasets (Brier score =0.070– 0.156). Consequently, we selected the stacking ensemble model for subsequent development. Subsequently, we compared the stacking ensemble-based radiomics model with the clinical model and the H/CL ratio for diagnosing CA. As shown in ***Table 2 and* *Figure 3***, the radiomics model achieved higher AUCs than the clinical model across all cohorts. A statistically significant difference was observed in this Center1-validation set between the radiomics model and the clinical model (AUC: 0.871 vs. 0.546, p = 0.01), whereas no significant differences were found in the other dataset. Compared with the H/CL ratio, the radiomics model yielded higher AUCs in all cohorts except Center 3, with a significant difference observed in the Center 1-training set (0.953 vs. 0.859, p = 0.023).

**Figure 2.**
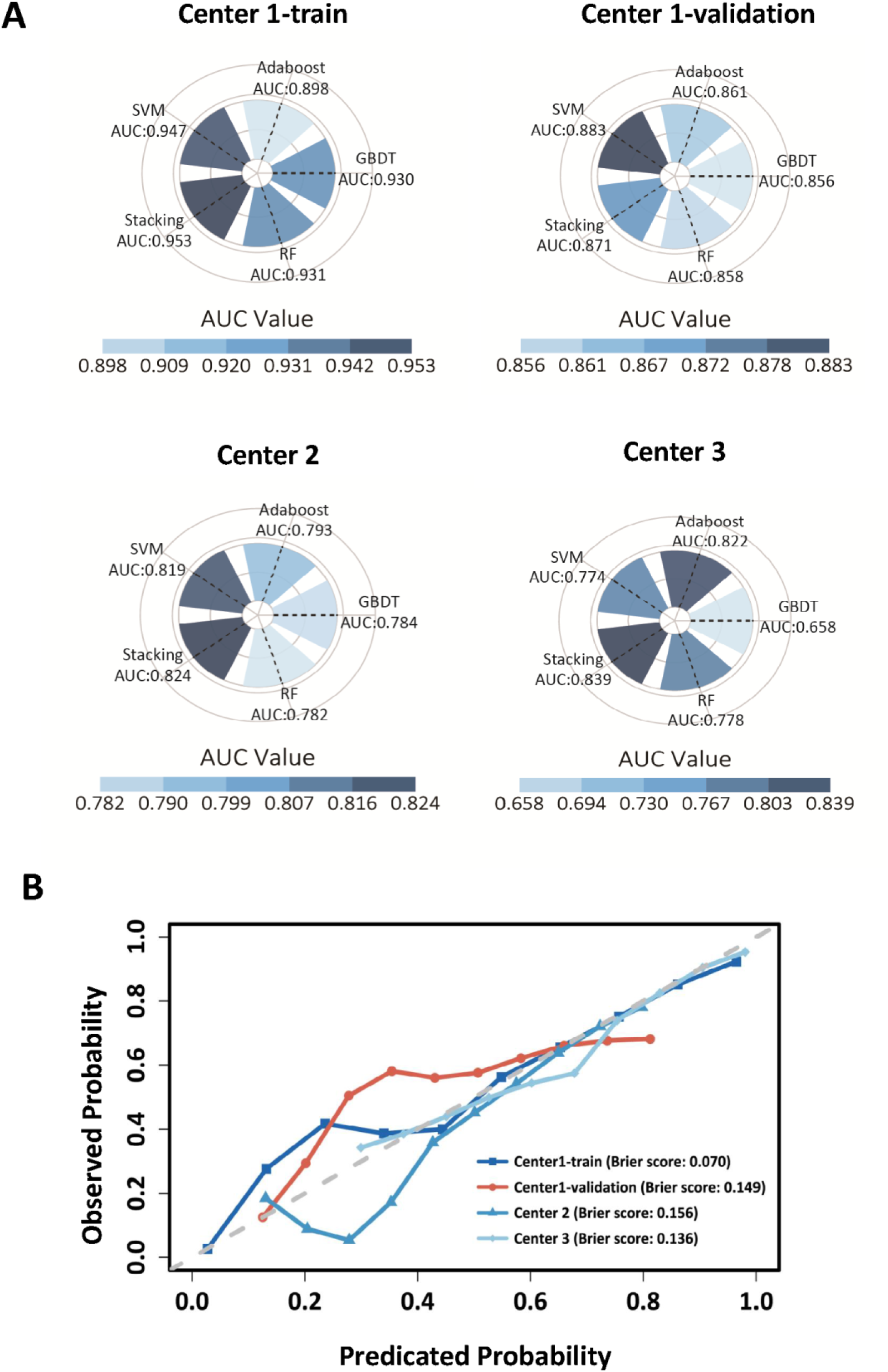
Performance comparison of machine learning models across multicenter cohorts using radar plots. (A) The radar charts display AUC values for five models: Support Vector Machine (SVM), Random Forest (RF), Gradient Boosting Decision Tree (GBDT), Adaptive Boosting (AdaBoost), and Stacking Ensemble, evaluated on Center 1 training set, Center 1 validation set, Center 2 test set, and Center 3 test set. (B) Calibration curves of the radiomics model for the Center 1 training set, Center 1 validation set, Center 2 test set and Center 3 test set.

**Figure 3.**
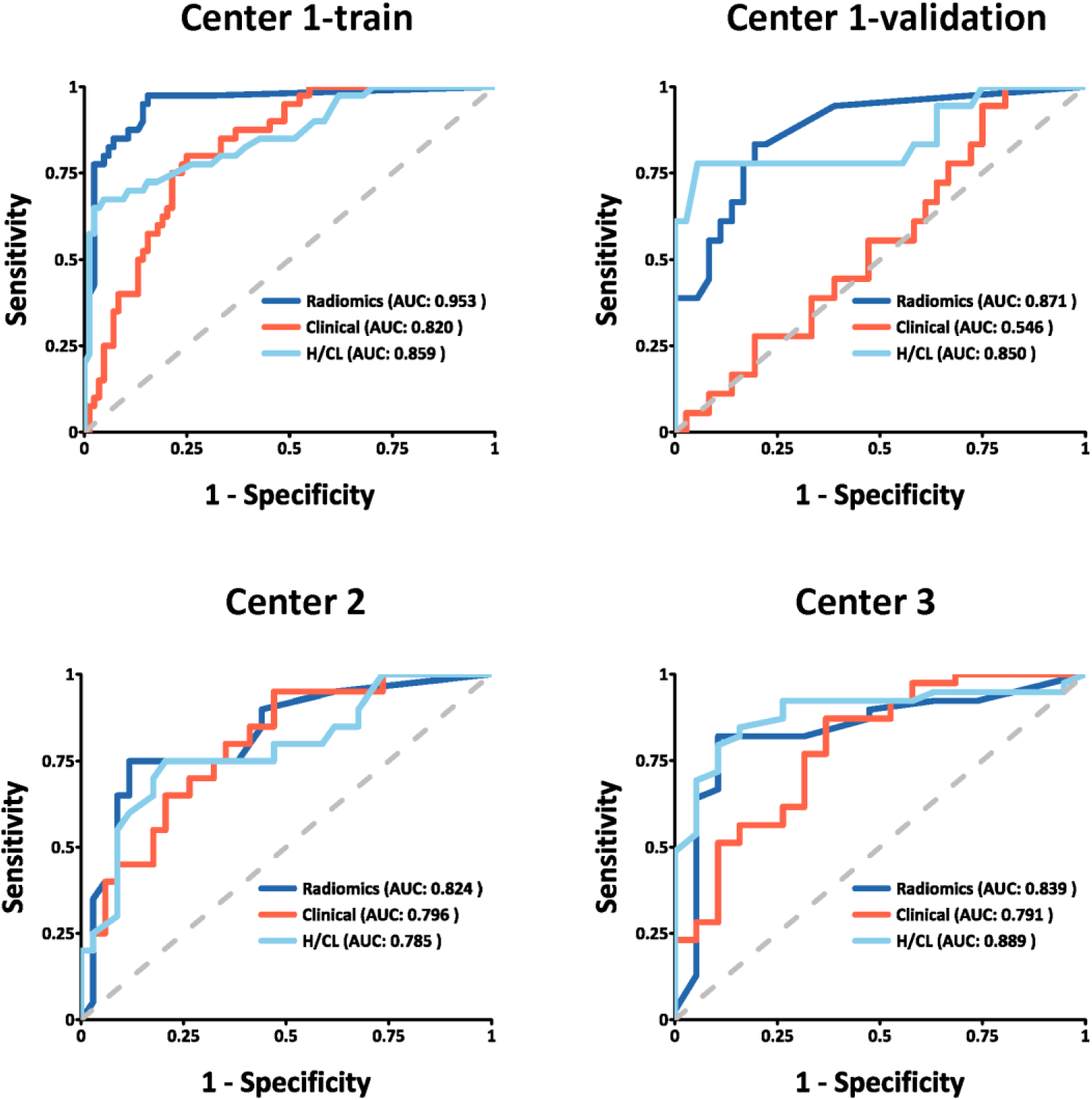
Receiver operating characteristic (ROC) curves of the Radiomics, Clinical, and H/CL models for distinguishing CA from non-CA. ROC curves are presented for Center 1 training set, Center 1 validation set, Center 2 test set, and Center 3 test set.

**Table 2.**
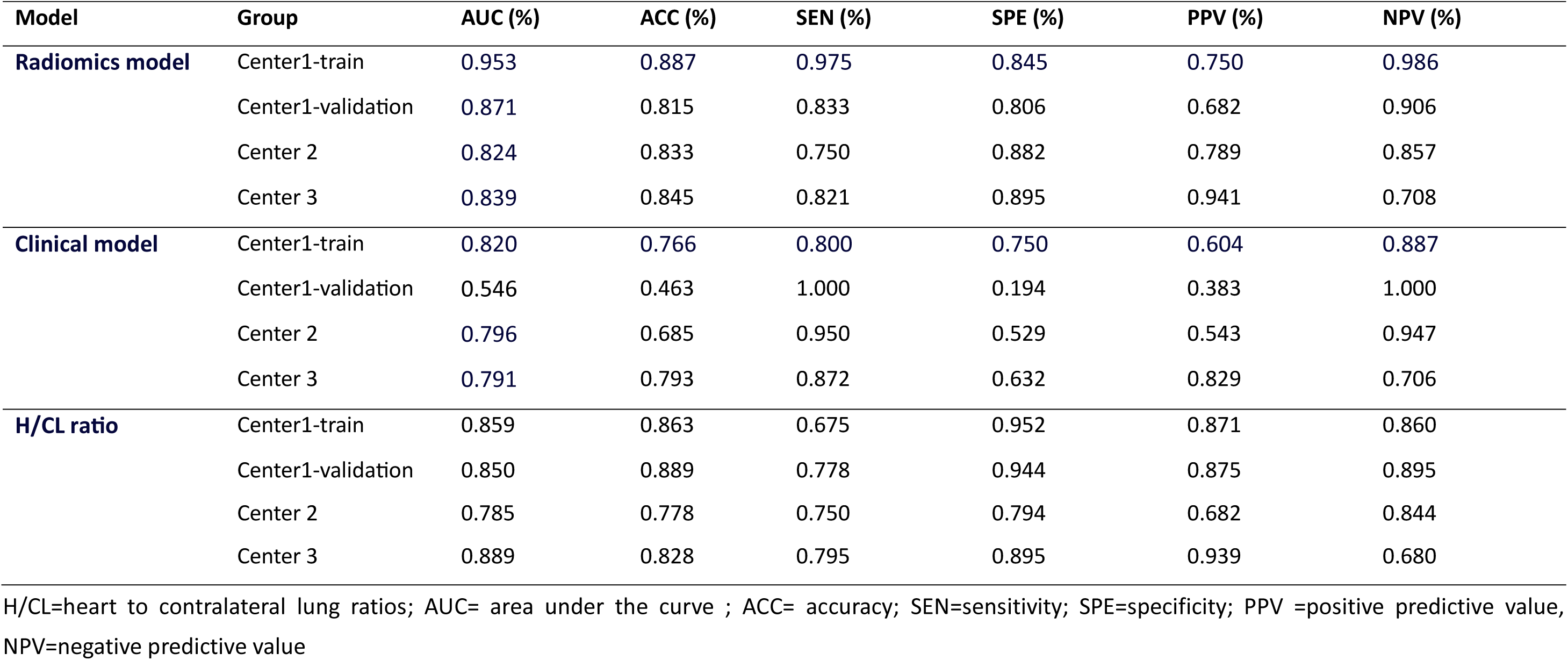
Diagnostic Performance of the SPECT/CT Radiomics Model, Clinincal Model and H/CL ratio among Different Cohorts.

DCA assessed the clinical utility of the predictive model by comparing net benefits at various threshold probabilities (***Figure 4***). The results demonstrated that the radiomics model consistently provided a higher net benefit than the clinical model across most risk thresholds, while its performance was comparable to that of the H/CL ratio.

**Figure 4.**
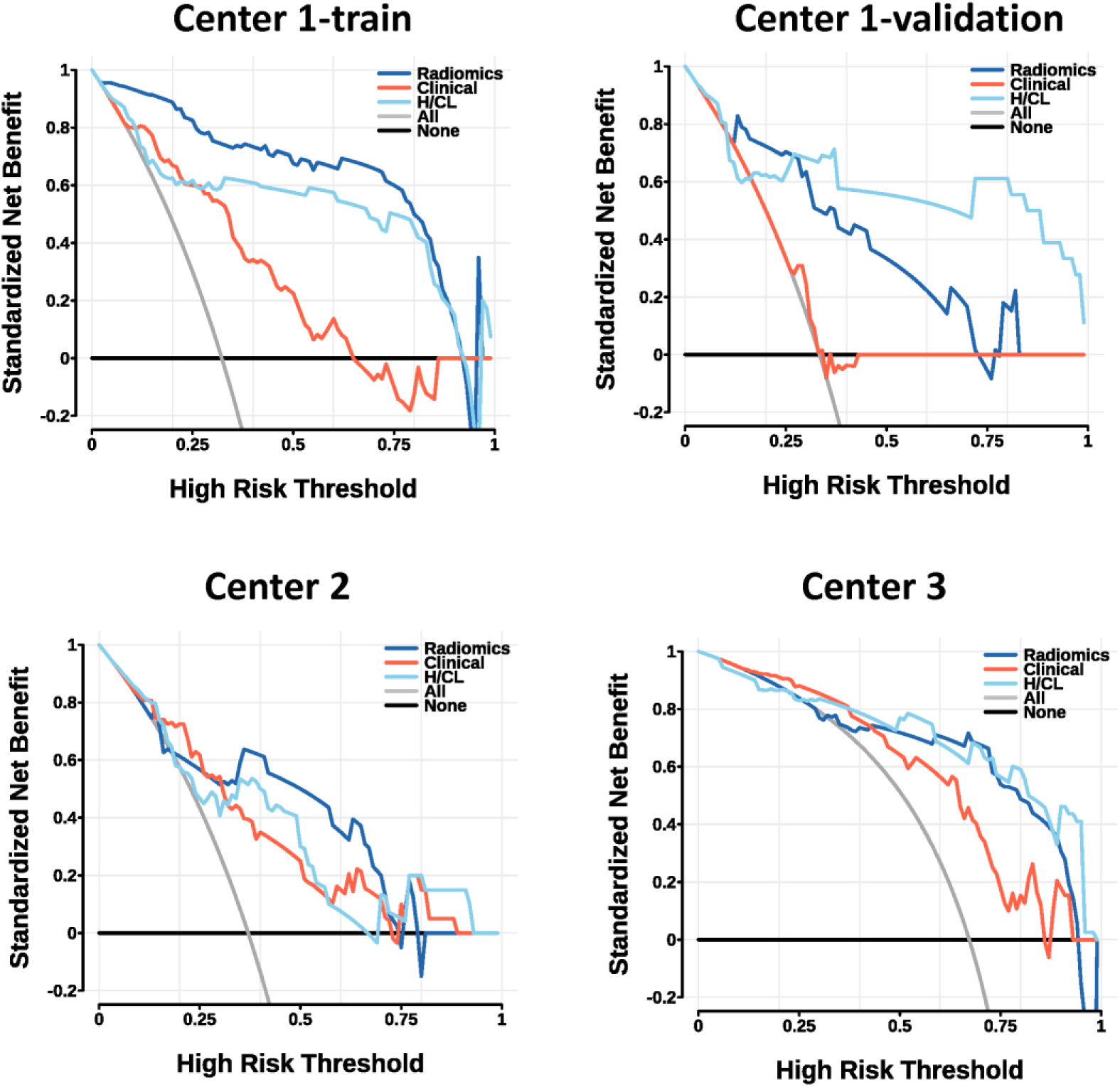
Decision curve analysis (DCA) of Radiomics, Clinical, and H/CL models. DCA curves are presented for Center 1 training set, Center 1 validation set, Center 2 test set, and Center 3 test set.

Figure 5 displays the SHAP summary plots for the top-performing models, highlighting both SPECT- and CT-derived features. These SHAP plots illustrate the relative contribution of each feature, offering valuable insights into the model’s decision-making process and enhancing interpretability of the ML predictions.

**Figure 5.**
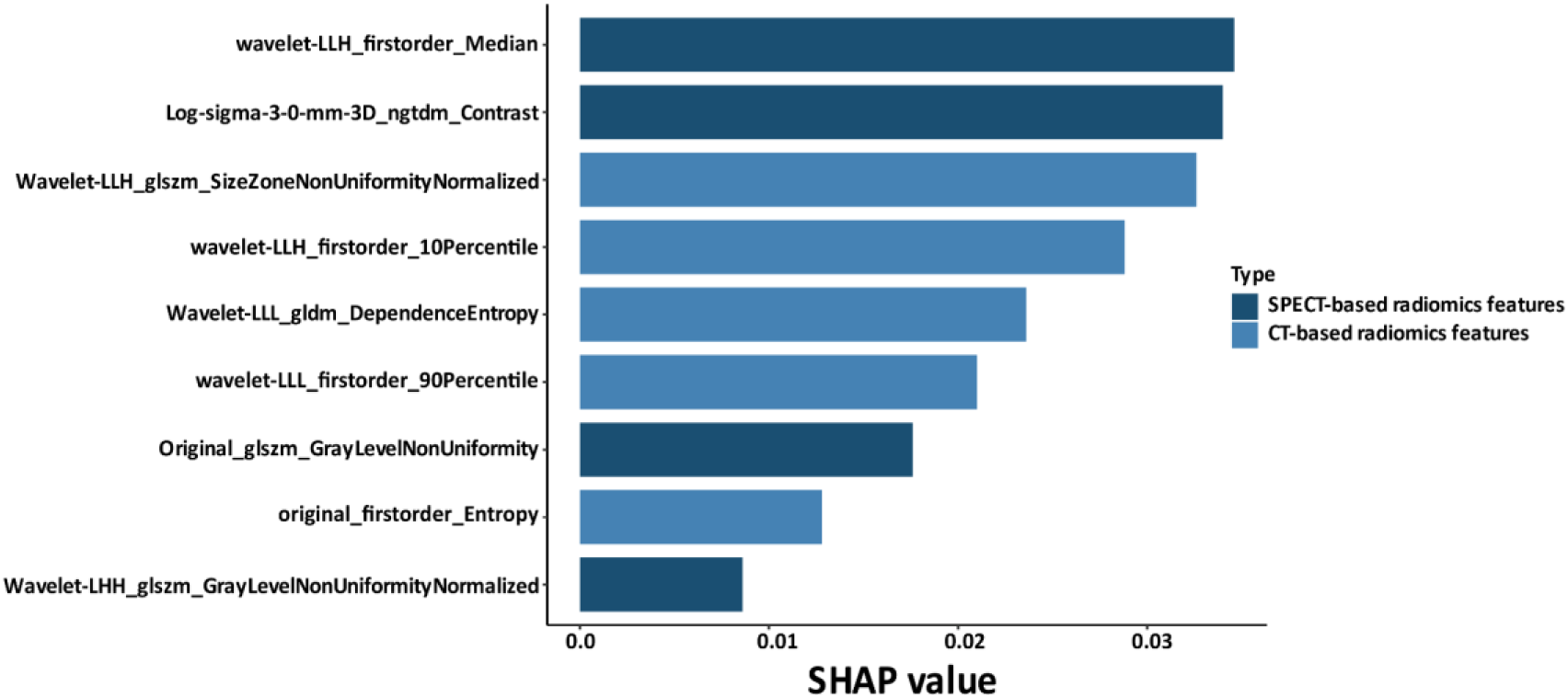
SHAP (SHapley Additive exPlanations) summary plot displaying the impact of various features for radiomics model.

### Model Performance for Differentiating ATTR-CA and AL-CA

To further evaluate the model’s capability to discriminate ATTR-CA from AL-CA, we conducted another validation using data from three centers. Owing to the limited number of CA cases in Centers 2 and 3, these cohorts were combined into a single subgroup. The established radiomics model was then applied independently to the Center 1 cohort and the merged Centers 2&3 cohort. As detailed in ***Supplementary Table 3***, the model achieved higher accuracy in distinguishing ATTR-CA from AL-CA than in differentiating CA from non-CA in both test sets.

## DISCUSSION

This multicenter study represents the first successful implementation of an ensemble ML model for automated CA diagnosis using hybrid SPECT/CT radiomics. Our key findings demonstrate that the stacking ensemble model integrating SVM, RF, GBDT, and AdaBoost achieved superior diagnostic performance compared to individual classifiers and clinical parameters, with robust generalizability across geographically distinct countries and tracers.

The current diagnostic process for CA is complex and time-consuming, highlighting the need for reliable and non-invasive tools to enable early and accurate detection[9, 20, 21]. Patients with CA often present with comorbidities such as increased NT-proBNP levels and left ventricular hypertrophy, which have been identified as predictive factors in prior studies[22-24]. Our research identified lgNT-proBNP and IVS thickness as independent risk factors for CA, consistent with existing literatures. This limited performance is likely due to the clinical overlap between CA and other cardiomyopathies, such as hypertrophic cardiomyopathy and hypertensive cardiomyopathy, which also present with heart failure with preserved ejection fraction (HFpEF) and left ventricular hypertrophy. Therefore, relying solely on these two markers lacks the specificity required for accurate CA diagnosis.

The use of [^99m^Tc]Tc-bisphosphonate imaging has become increasingly widespread due to its high diagnostic accuracy and growing awareness of ATTR-CA[25]. For example, Perugini grading of planar [^99m^Tc]Tc-bisphosphonate scans can reach very high sensitivity (99%) for ATTR-CA but at the cost of reduced specificity (86%) due largely to false positives in AL-CA[26]. Similarly, the commonly used H/CL ratio threshold (≥1.3 at 3 h) helps confirm ATTR-CA when uptake is equivocal[26], but its added value over expert reading is modest. In contrast, our model’s automated quantitative analysis consistently outperformed these traditional metrics.

Notably, the model’s diagnostic accuracy matched or exceeded that of previously reported AI approaches. For instance, a ML model based on clinical information, laboratory tests, and imaging examinations achieved an AUC of 0.84 in detecting ATTR-CA among patients with AS[7]. A coronary CT-derived radiomics model achieved AUC 0.91 for CA[27], while a deep-learning pipeline for SPECT/CT quantitation reported A near-perfect AUC (0.99) in distinguishing ATTR-positive scans[4]. Likewise, other AI approaches to bone scintigraphy have reported AUCs in the 0.88-0.94 range[28]. Moreover, the results showed that the diagnostic performance of our radiomics model in the Austrian cohort using [^99m^Tc]Tc-DPD (AUC: 0.839) was comparable to that in the Chinese cohort using [^99m^Tc]Tc-PYP (AUC: 0.824). This consistency across tracers suggests that the model’s performance is largely independent of the specific radiopharmaceutical used. Taken together, these comparisons suggest that our hybrid SPECT/CT radiomics model provides enhanced sensitivity and specificity for CA, independent of imaging protocols and tracers.

The improved accuracy is likely attributable to the rich information captured by combining SPECT and CT features. Radiomics analysis extracts subtle patterns of tracer uptake heterogeneity in the SPECT data alongside texture and density features from the CT images. Each modality offers complementary clues: SPECT reflects the degree and distribution of amyloid-binding tracer activity, while CT encodes structural changes (e.g. myocardial thickening, fibrosis). Our SHAP analysis identified wavelet-transformed first-order and texture features as the most influential predictors, reflecting the distribution of pixel intensities within specific frequency bands. This highlights their significance in quantitative SPECT/CT radiomics and their ability to capture subtle pathophysiological information that escapes visual detection. Importantly, our model also demonstrated the ability to classify amyloid subtype. It distinguished ATTR from AL with high accuracy, even outperforming the CA vs. non-CA task. This likely reflects known tracer-binding biology: ATTR-amyloid avidly takes up bone-avid tracers, whereas AL-amyloid shows little or no uptake. In practice, Perugini Grade ≥2 uptake without monoclonal proteins virtually confirms ATTR-CA. Our radiomics model learned these distinctions automatically, capturing subtle texture features linked to tracer retention. Unlike prior AI tools focused only on detecting amyloid presence, our approach enables subtype differentiation-critical for guiding therapy, as ATTR and AL require fundamentally different treatments (stabilizer drugs vs. chemotherapy).

These findings have important clinical implications. Early and accurate CA diagnosis is crucial because effective treatments now exist, especially for ATTR (e.g. tafamidis) and AL (chemotherapy, stem cell transplant). Our radiomics model could flag patients who might otherwise be missed by visual readout, enabling prompt intervention. For example, AI-enhanced scintigraphy might detect low-grade tracer uptake or atypical patterns, prompting further workup. By reducing equivocal results, the model may also decrease the need for invasive procedures like biopsy. Moreover, as many CA patients are initially misdiagnosed or diagnosed late, our approach holds promise for earlier detection. In sum, integrating such a model into clinical imaging could enhance confidence in non-invasive diagnosis, guide appropriate workup (e.g. CMR, genetic testing), and ultimately improve patient outcomes.

This study has several limitations. (1) As a retrospective study, it is inherently prone to selection bias, which is difficult to avoid. (2) The sample size of this study is relatively small, and the results need to be validated in a multicenter study with a larger sample size. (3) Although we intentionally included data from three centers and multiple scanner types to improve generalizability, device-dependent variability in image texture and reconstruction could still influence performance. In particular, the relative scarcity of AL-CA cases in our training set may have introduced class imbalance, this could lead the model to favor ATTR-like patterns; (4) Like most imaging AI studies to date, our work relied on existing labeled data and did not include prospective validation.

## CONCLUSION

This study establishes that ensemble ML model of SPECT/CT radiomics significantly outperforms conventional diagnostic metrics for CA detection, demonstrating multicenter robustness and clinically actionable calibration, supporting its value in early CA diagnosis and clinical decision-making.

## Statements & Declarations Fundings

This work was funded by National Natural Science Foundation of China (No. 82472029 and 81901784 to M.Z.).

## Competing interests

C.P.S. reports speaker and consulting fees from Pfizer. All other authors have no relationships relevant to the contents of this paper to disclose.

## Author Contributions

Q.M and M.Z contributed to the study design. Material preparation, data collection and analysis were performed by Q.M, J.C, S.J, Y.Z, Y.X, C.L, C.Z and C.P.S. The first draft of the manuscript was written by Q.M and reviewed by R.C,K.C, M.H, X.L, W.Z and M.Z. All authors commented on previous versions of the manuscript. All authors read and approved the final manuscript.

## Data availability

The data underlying this article will be shared on reasonable request to the corresponding author.

## Ethics approval

This study was granted ethical approval by the institutional review boards of Xiangya Hospital, Central South University (No. 202310202), Fuwai Central China Cardiovascular Hospital (Ethics Review No. 202303), and the General Hospital of Vienna (EK1557/2020). It was carried out in accordance with the principles of the 1964 Declaration of Helsinki. Owing to the retrospective study design, the Institutional Review Boards of all participating institutions formally waived the requirement for informed consent.

## Data Availability

The data underlying this article will be shared on reasonable request to the corresponding author.

## Notes

### Author Declarations

This study was granted ethical approval by the institutional review boards of Xiangya Hospital, Central South University (No. 202310202), Fuwai Central China Cardiovascular Hospital （Ethics Review No. 202303), and the General Hospital of Vienna (EK1557/2020). It was carried out in accordance with the principles of the 1964 Declaration of Helsinki. Owing to the retrospective study design, the Institutional Review Boards of all participating institutions formally waived the requirement for informed consent.

