## Supplementary Data for "Stacking Ensemble Learning-based Models Enabling Accurate Diagnosis of Cardiac Amyloidosis using SPECT/CT:an International and Multicentre Study"

**Supplementary Table 1.** SPECT/CT Acquisition Parameters by Center

| | **Center 1** | **Center 2** | **Center 3** |
|---|---|---|---|
| SPECT/CT camera | Philips Precedence16 | GE 870 DR | Siemens Symbia |
| **SPECT parameters** | | | |
| Reconstruction algorithm | ASTONISH | OSEM | 3DOSEM |
| Energy window | 140 keV, 20% | 140 keV, 20% | 141 keV, 15% |
| Matrix size | 64×64 | 256×256 | 256×256 |
| Number of views | 64 | 40 | 64 |
| Time per stop | 25 seconds | 25 seconds | 20 seconds |
| SPECT slice thickness | 9.3 mm | 2.2 mm | 2.4 mm |
| **CT parameters** | | | |
| CT Voltage (kV) | 120 | 120 | 130 |
| CT Tube current (mAs) | 180 | 260 | 80 |
| CT slice thickness | 3.3 mm | 3.8 mm | 3.0 mm |

CT=Computed Tomography; SPECT=Single Photon Emission Computed Tomography

**Supplementary Table 2.** Univariate and multivariable logistic regression analysis of clinical factors on the training set

| | **Univariate logistic analysis** | | **Multivariate logistic analysis** | |
|---|---|---|---|---|
| | OR (95%CIs) | P-value | OR (95%CIs) | P-value |
| Age, per 1 year | 1.04 (1.01 -1.08) | 0.016 | 1.02 (0.98 - 1.06) | 0.216 |
| Male | 0.83 (0.35 -1.97) | 0.677 | | |
| Diabetes | 0.24 (0.05 -1.12) | | | |
| Hypertension | 0.73 (0.33 -1.64) | 0.451 | | |
| NYHA class ≥III | 1.48 (0.68 -3.20) | 0.322 | | |
| lgNT-proBNP, per 1 ng/mL | 5.31(2.48-11.37) | <0.001 | 4.63 (2.09 - 10.29) | <0.001 |
| eGFR, per 1mL/min/1.73m$^2$ | 1.00 (1.00 - 1.01) | 0.242 | | |
| IVS thickness, per 1 mm | 1.25 (1.10 - 1.42) | <0.001 | 1.23 (1.07 - 1.40) | 0.003 |
| LVEF, per 1 % | 1.00 (0.99 - 1.01) | 0.518 | | |

OR=odds ratio; CI=confidence Interval; NYHA=New York Heart Association; NT-proBNP= N-terminal of the prohormone brain natriuretic peptide; eGFR=estimated glomerular filtration rate; IVS=Interventricular septal; LVEF=Left ventricular ejection fraction

**Supplementary Table 3.** Diagnostic Performance of the SPECT/CT Radiomics Model among Different Cohorts

| | AUC (%) | ACC (%) | SEN (%) | SPE (%) | PPV (%) | NPV (%) |
|---|---|---|---|---|---|---|
| **ATTR-CA vs. AL-CA** | | | | | | |
| Center 1 (n=53) | 0.989 | 0.962 | 0.947 | 1.000 | 1.000 | 0.882 |
| Center 2&3 (n=55) | 0.904 | 0.873 | 0.842 | 0.941 | 0.970 | 0.727 |
| **CA vs. non-CA** | | | | | | |
| Center 1 (n=178) | 0.918 | 0.888 | 0.879 | 0.892 | 0.797 | 0.939 |
| Center 2&3 (n=112) | 0.823 | 0.795 | 0.712 | 0.887 | 0.875 | 0.734 |

CA= CA=cardiac amyloidosis; ATTR-CA= transthyretin cardiac amyloidosis； AL-CA=
AUC= area under the curve ; ACC= accuracy; SEN=sensitivity; SPE=specificity; PPV =positive predictive value, NPV=negative predictive value

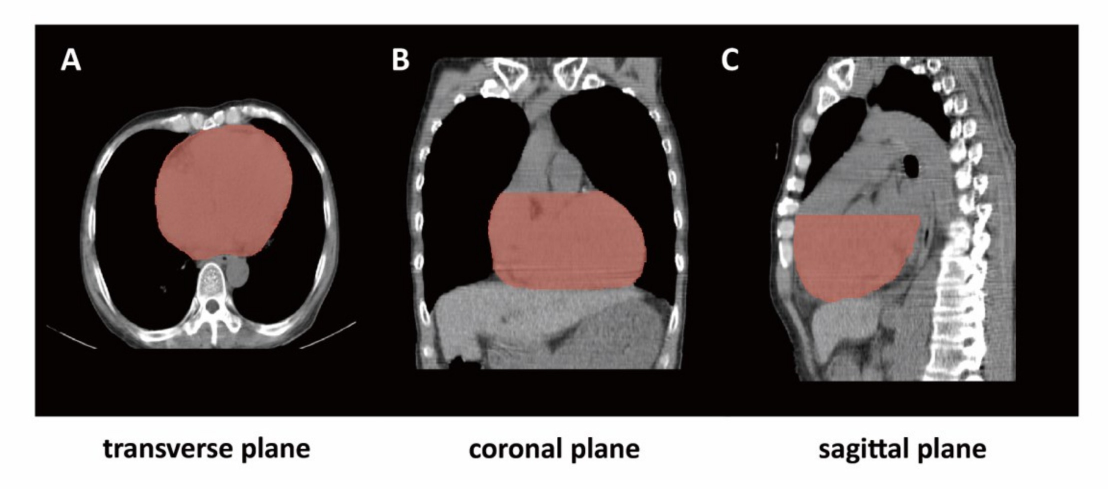


**Supplementary Fig 1.** Automatically delineated volume of interest (VOI) of the heart. (A) The transverse plane, (B) the coronal plane and (C) the sagittal plane.

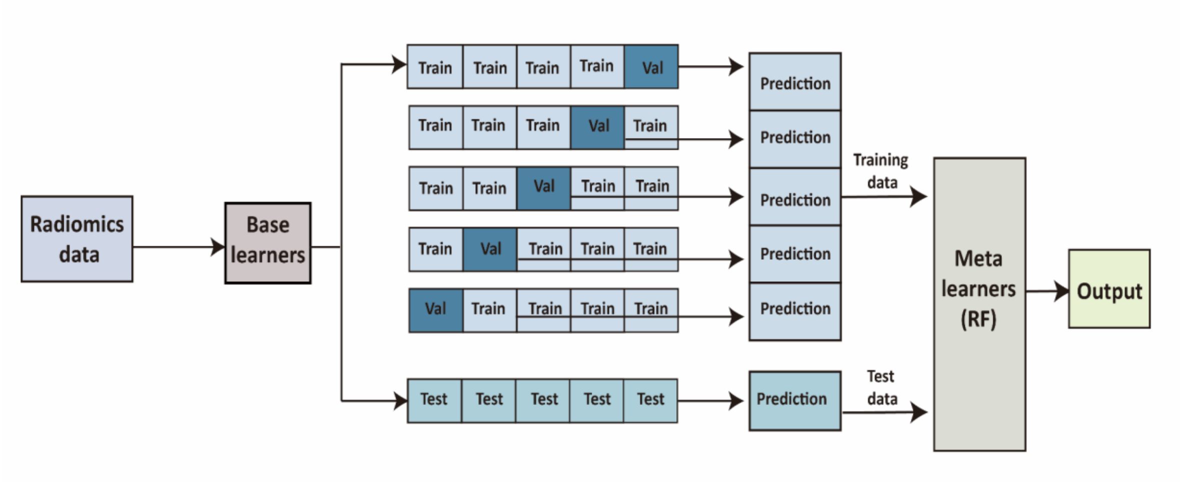


**Supplementary Fig 2.** The development process of the stacking model. The stacking model employed Adaptive Boosting (AdaBoost), Gradient Boosting Decision Tree (GBDT), Random Forest (RF), and Support Vector Machine (SVM) as base learners (first-level), with RF serving as the meta-learner (second-level). To prevent data overfitting, a 5-fold cross-validation strategy was applied to the training set of base learners. Specifically, the original training data were partitioned into 5 subsets. In each iteration, four subsets were utilized to train these four base learners, while the remaining subset served as the validation set to generate predicted probability values. After five iterations, each subset had obtained predicted probability values. All probability predictions from the base learners were aggregated into a new feature matrix, which constituted the meta-training set for the second-level learn
